# Performance of upper-arm capillary blood collection for Alzheimer’s disease and central nervous system biomarkers: comparison of Tasso+ and venous plasma

**DOI:** 10.64898/2026.08.13.26360406

**Authors:** Taraneh E. Atri, Marisa N. Denkinger, James Liu, Alpana Singh, Michelle Surdyn, Victoria A. Brown, Guadalupe Martinez, Marlene Teran, Vanessa Soza, Angela Kuramoto, Tainá M. Marques, Jessica B. Langbaum, Alireza Atri, Nicholas J. Ashton

## Abstract

**INTRODUCTION:** Novel capillary-blood collection methods have not yet been evaluated for a wide range of central nervous system (CNS) and neurodegenerative disease-related proteins. Biomarkers of Alzheimer’s disease (AD) and related disorders (ADRD) collected from devices like the Tasso+, a minimally invasive upper-arm capillary blood collection device, must be compared to traditional venipuncture to assess for validity.

**METHODS:** Participants underwent blood collection via traditional venipuncture and Tasso+ in a clinical research setting. The Nucleic Acid Linked Immuno-Sandwich Assay (NULISA) CNS panel was used for biomarker quantification in venous and Tasso-derived plasma.

**RESULTS:** Eighty-three participants (age mean±SD 76.8±8.2 years, 79.5% cognitively unimpaired) completed blood collection. Little to no correlation was found between venous and Tasso+ plasma for p-tau217, but the correlation was improved by using a brain-derived (BD)-p-tau217/BD-p-tau181 ratio. Extremely strong correlations were found for neurofilament light (NfL) and glial fibrillary acidic protein (GFAP). Among the 131 biomarkers measured, 51 (38.9%) had a Pearson R ≥ 0.90; 27 (20.6%) had values between 0.70-0.90; 26 (19.9%) had values between 0.30-0.70; and 27 (20.6%) had values ≤0.30.

**DISCUSSION:** The Tasso+ accurately measures NfL and GFAP, but caution is warranted when measuring other AD/ADRD biomarkers, as agreement with venous plasma appears to be protein or ratio dependent. These results highlight that important biomarker-specific differences must be considered when translating capillary blood collection approaches. They also further support foundations for development of these methods, highlighting both the opportunities and remaining challenges for translating the promise of blood-based biomarkers beyond AD/ADRD specialty clinics and research settings.

## 1. Introduction

Blood biomarkers that reflect the pathophysiology of Alzheimer’s disease (AD) and related disorders (ADRD) are valuable tools in research, therapeutic trials, and increasingly, clinical care [1]. Phosphorylated tau at threonine 217 (p-tau217), either alone or in combination with amyloid-β42 (Aβ42), has emerged as the leading Core 1 diagnostic blood biomarker for AD, accurately predicting amyloid-β plaque pathology measured by positron emission tomography (PET), to support amyloid “A” status in the ATX(N) AD classification [2,3]. Additionally, neurofilament light (NfL) and glial fibrillary acidic protein (GFAP) reflect neurogenerative and neuroinflammatory processes, respectively, providing supporting information for non-specific AD processes and have broader uses across the neurodegenerative and ADRD spectrum [4–9].

Currently, blood biomarkers such as plasma p-tau217 are primarily used in specialized memory clinics during evaluation of symptomatic individuals with suspected AD. However, such biomarkers could be implemented across broad, unselected, and diverse populations. Such approaches could facilitate more accurate global or community-specific AD prevalence estimates, identification of eligible individuals for prevention trials and, possibly soon, for clinical care [10–13]. Their integration into primary care and community-based settings would likely reduce testing barriers, support more efficient and equitable biomarker-guided care and research participation and improve representation of historically underserved high risk AD populations. Population-level and real-world primary care clinic AD/ADRD blood biomarker data that fully captures the diversity of unselected populations are still scarce. One contributing key factor is that current AD blood biomarker testing relies on venipuncture and requires standardized post-collection processing procedures, laboratory resources, and competence. This introduces logistical challenges including the need for phlebotomists, immediate sample processing, and cold-chain shipping [14].

Given the challenges associated with venipuncture, recent advances in blood collection have increasingly focused on minimally invasive alternatives, including capillary blood sampling from the upper arm or fingertip, with the latter collected onto dried blood spot (DBS) cards [15,16]. In this study, we evaluated the Tasso+ upper-arm capillary blood collection device because it was previously identified as the least painful and most preferred sampling method in a comparative study of minimally invasive collection devices [17]. Previous studies have also demonstrated excellent agreement between Tasso-derived plasma or serum and conventional venous plasma for a range of peripheral analytes, including albumin, chloride, magnesium, testosterone and glucose [18,19]. While these successful studies provide valuable information, biomarkers for AD/ADRD and neurodegenerative diseases have not yet been widely evaluated in upper arm capillary blood.

In this study, we investigated central nervous system (CNS)-derived proteins in Tasso+ plasma focusing on biomarkers relevant to AD and related neurodegenerative disorders. Biomarker measurements were performed using the Nucleic Acid Linked Immuno-Sandwich Assay (NULISA™) CNS Disease Panel 120 (Alamar Biosciences), which quantifies 131 CNS- and neurodegeneration-related proteins from small volumes of biofluids [20,21]. The panel includes established AD biomarkers, such as p-tau217, brain-derived p-tau217 (BD-p-tau217), NfL, and GFAP, alongside a range of additional proteins implicated in neurodegeneration, synaptic dysfunction, neuroinflammation, and other CNS pathways. By comparing biomarker concentrations in Tasso-derived and venous plasma, we aimed to evaluate the suitability of upper-arm capillary blood collection for measuring established and emerging AD/ADRD-related blood biomarkers.

## 2. Methods

### 2.1 Participants and Sample Collection

#### 2.1.1 Consent Statement

A total of 87 participants were recruited from longitudinal cohort studies of aging and neurodegenerative disorders and the Memory/Cognitive Disorders clinic at Banner Sun Health Research Institute (Sun City, Arizona).

All participants’ cognitive, functional and clinical status were characterized through integrated assessment of validated instruments and cognitive testing. Participants underwent informed consent process and assessment of decisional capacity and they, or their legally authorized representatives, as appropriate, provided signed consent. The study was approved by the WCG Western Institutional Review Board.

#### 2.1.2 Sample collection

Each participant’s capillary blood was collected using the Tasso+ and venous blood was collected by standard venipuncture. Both sample types were collected into ethylenediaminetetraacetic acid (EDTA) anticoagulant tubes. For Tasso+ collection, the upper arm was warmed with a heat pack then cleansed with 70% isopropyl alcohol. The device was applied to the upper arm and activated according to the manufacturer’s instructions by pressing the release button to deploy the lancet. Blood was allowed to collect for up to 10 minutes or until the collection tube was filled. Following collection, the tube was capped and gently inverted 10–12 times to ensure adequate mixing with the anticoagulant. A phlebotomist collected venous blood into 10 mL EDTA tubes, which were similarly inverted 10–12 times immediately after collection. All samples were transferred directly to laboratory personnel for processing immediately after acquisition.

### 2.2 Sample Processing

Samples were immediately placed on wet ice. Venipuncture tubes were centrifuged at 3500×g for 10 minutes at 4°C. Venous plasma was then aliquoted into 1.5 mL protein LoBind tubes (Eppendorf). Tasso+ EDTA tubes were also centrifuged at 3500×g for 10 minutes at 4°C. Plasma was transferred to a 1.5 mL protein LoBind tube and re-spun with the same settings. Tasso+ plasma was then aliquoted into a final 1.5 mL protein LoBind tube. Samples were stored at −80°C until analysis.

### 2.3 Plasma biomarker analysis

On the day of analysis, plasma aliquots from venous and Tasso-derived blood draws were thawed at room temperature. For NULISA, plasma samples were spun at 10,000×g for 10 minutes at 4°C. A total of 25 µL were then loaded onto an Alamar-provided 96-well plate. DNA reporter outputs were pooled to form libraries which were amplified through polymerase chain reaction (PCR) and sequenced in-house using the AVITI 2×75 Cloudbreak Freestyle High Output Kit on the AVITI sequencer (Element Biosciences). Venous plasma samples were also analyzed on the cobas e801 instrument (Roche Diagnostics) with the Elecsys pT217 RUO assay to determine if amyloid pathology was present in recruited participants based on defined cut-offs. Cut-offs were defined using a cohort of 616 participants from the Wisconsin Registry for Alzheimer’s Prevention (WRAP) and are based on prediction of amyloid-PET positivity using a 24.1 centiloid cutoff.

### 2.4 Data processing and normalization

FASTQ DNA files generated by the sequencer were uploaded to the NULISA analysis software (Alamar Biosciences) for processing using the NULISAseq algorithm. Both intraplate and interplate normalization were performed using internal control samples. Lastly, data was rescaled and log2-transformed to yield NULISA Protein Quantification (NPQ) units.

### 2.5 Statistical analysis

All statistical analyses and plots were created using RStudio with R v4.5.2 (10-31-2025). Linear regression models were used to determine trends, and 95% confidence intervals were determined using the standard error. Correlations were tested using the Pearson correlation coefficient (*R*) and nominal p values were reported.

## 3. Results

### 3.1 Cohort Demographics

Eighty-three of 87 recruited participants were included in the analysis. Four participants were excluded due to insufficient Tasso+ plasma volume at collection (<20 µL). Included participants had mean age 76.8 ± 8.2 years, were 68.7% female, 66 (79.5%) were cognitively unimpaired (CU), 9 (10.9%) had Mild Cognitive Impairment (MCI), and 8 (9.6%) were diagnosed with dementia due to AD or a related neurodegenerative condition (AD/Dementia) (**Table 1**). Overall, 36.1% of participants had high p-tau217 levels (≥0.326 pg/mL), 26.5% had intermediate levels, and 37.4% had low levels (≤0.201 pg/mL) as determined by the Elecsys pT217 RUO assay.

**Table 1.**
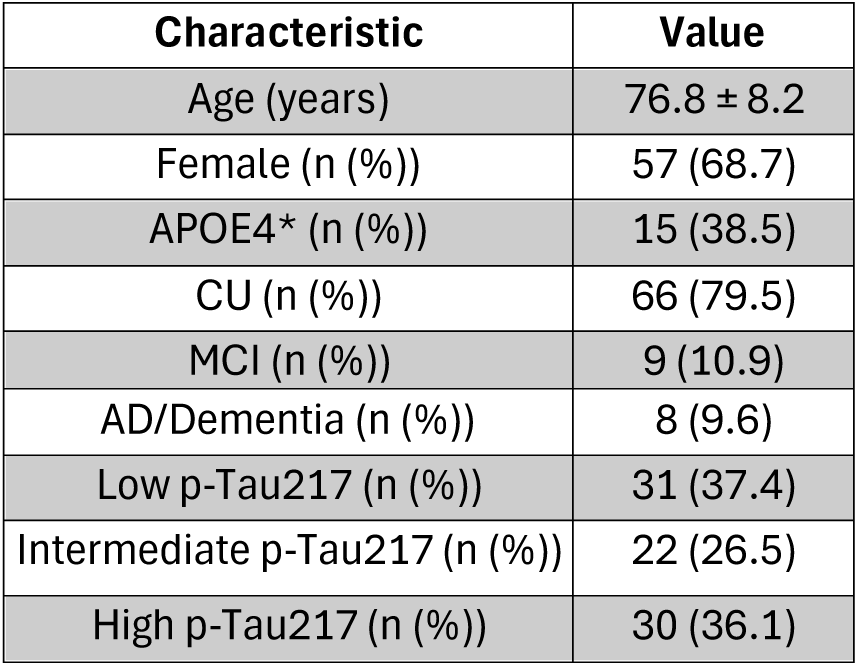
Demographics table for the 83 participants included in analysis. *APOE4 value represents those that have one or more *APOE4* genes.

| Characteristic | Value |
| --- | --- |
| Age (years) | 76.8 ± 8.2 |
| Female (n (%)) | 57 (68.7) |
| APOE4* (n (%)) | 15 (38.5) |
| CU (n (%)) | 66 (79.5) |
| MCI (n (%)) | 9 (10.9) |
| AD/Dementia (n (%)) | 8 (9.6) |
| Low p-Tau217 (n (%)) | 31 (37.4) |
| Intermediate p-Tau217 (n (%)) | 22 (26.5) |
| High p-Tau217 (n (%)) | 30 (36.1) |

### 3.2 p-Tau and Aβ Single Biomarker Correlations

We found a weak correlation between venous and Tasso+ plasma for all forms of NULISA p-tau: p-tau217 (**Figure 1A**; *R=*0.28, p=0.011), p-tau181 (*R=*0.17, p=0.124), and p-tau231 (*R=*0.18, p=0.10). A weak correlation was also observed with Roche RUO p-tau217 (*R=*0.029, p=0.0079). Similarly, there were weak but significant relationships for brain-derived (BD) p-tau181 (*R=*0.23, p=0.038) and BD-p-tau231 (*R=*0.22, p=0.047). However, for BD-p-tau217 there was a moderate but significant correlation between venous and Tasso+ plasma (**Figure 1B**; *R=*0.53, p=3.5×10^-7^). Notably, the NPQ values, and thus relative concentrations, for all p-tau isoforms in Tasso+ plasma were consistently higher than those from venous plasma (mean [variance] NPQ: 13.29 [0.63] Tasso+ vs 10.95 [0.73] venous, p=1.72×10^-41^). There was a weak but significant correlation for both Aβ40 (*R=*0.35, p=0.001) and Aβ42 (*R=*0.43, p=6.1×10^-5^).

**Figure 1.**
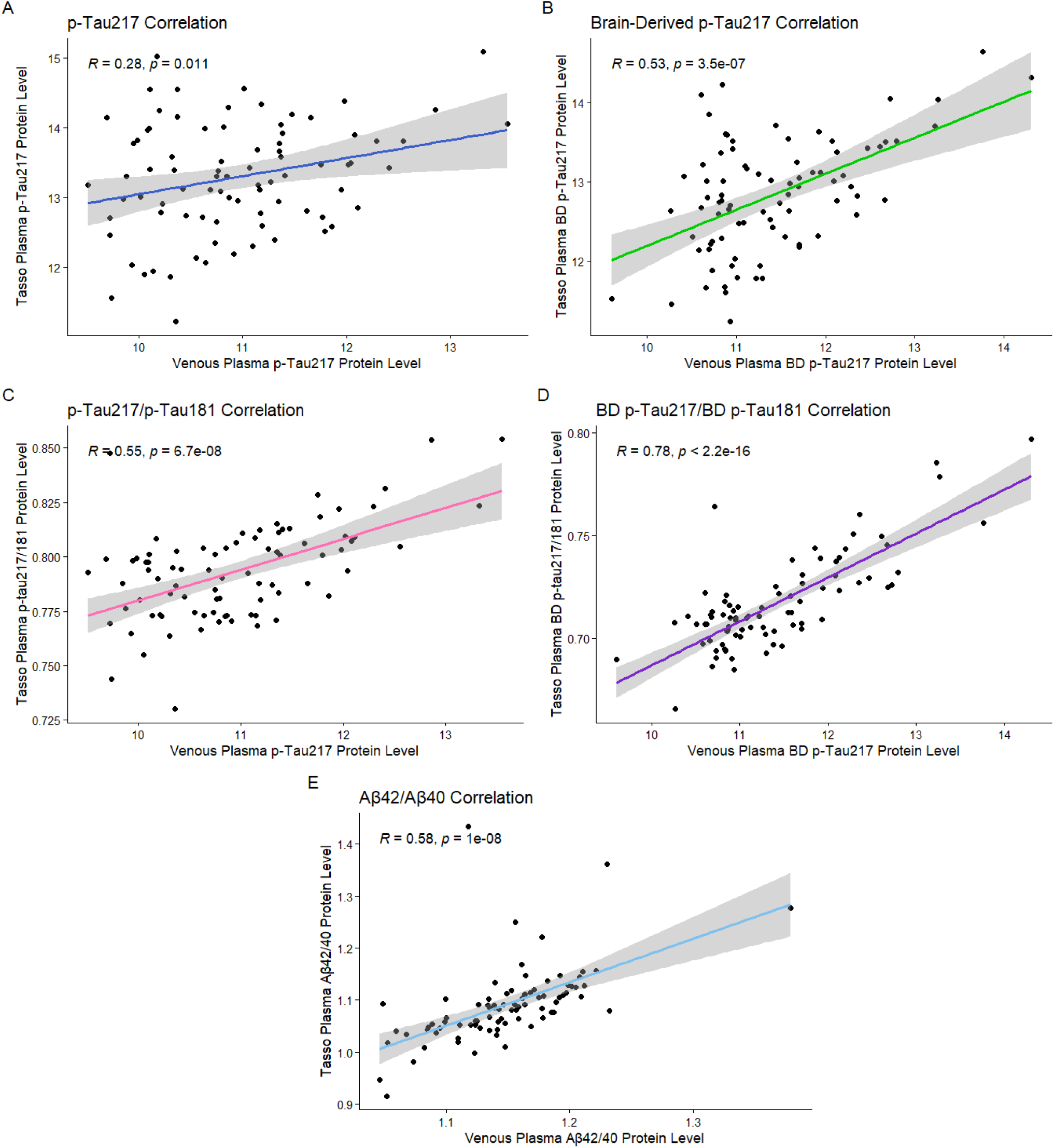
Biomarker correlations between venous and Tasso+ plasma for key Alzheimer’s disease biomarkers and ratios: p-tau217 (A), brain-derived p-tau217 (B), p-tau217/p-tau181 (C), BD p-tau217/BD p-tau181 (D), and Aβ42/40 (E). Lines represent linear regression results for Pearson’s R with 95% confidence intervals.

### 3.3 AD Biomarker Ratio Comparisons

To see if single biomarker correlations could be improved, ratios of biomarkers were tested in Tasso-derived plasma. Firstly, the ratio of p-tau217 and BD-p-tau217 to all other biomarkers was compared to venous p-tau217 and BD-p-tau217, respectively. Using the ratio of p-tau217 to p-tau181 (p-tau217/181) significantly improved the correlation strength when compared to that for the single-biomarker. (**Figure 1C**; *R=*0.55, p=6.7×10^-8^). However, the ratio of BD-p-tau217 to BD-p-tau181 (BD p-tau217/181) performed even better and yielded a strong correlation with venous BD-p-tau217 (**Figure 1D**; *R=*0.78, p<2.2×10^-16^). Finally, a moderate correlation was observed for the Aβ42/Aβ40 ratio (**Figure 1E**; *R=*0.58, p=1.0×10^-8^) in Tasso+ plasma when compared to the corresponding ratio in venous plasma.

### 3.4 All CNS Biomarker Results

While relationships observed for AD single biomarkers were not particularly strong, several other neurodegeneration biomarkers showed very strong correlations between venous and Tasso+ plasma. NfL (**Figure 2A**; *R=*0.96, p<2.2×10^-16^), NfH (**Figure 2B**; *R*=1.0, p<2.2×10^-16^), and GFAP (**Figure 2C**; *R=*0.95, p<2.2×10^-16^) all showed high concordance with similar absolute values. Among the 131 biomarkers measured in Tasso+ and venous plasma, 51 (38.9%) had a Pearson R greater or equal to 0.90; 27 (20.6%) had values between 0.70 and 0.90; 26 (19.9%) had values between 0.30 and 0.70; and 27 (20.6%) had values less than 0.30 (**Figure 3**). The five biomarkers with the lowest correlations are: TAR DNA-binding protein 43 (TARDBP, *R=*-0.0002), alpha-synuclein (SNCA, *R=*-0.005), oligodendrocyte alpha-synuclein (Oligo-SNCA, *R=*0.014), superoxide dismutase 1 (SOD1, *R=*0.018), and brain-derived neurotrophic factor (BDNF, *R=*0.024). Notably, most of the biomarkers, and especially those with low correlations, had relatively higher NPQs/concentrations in Tasso+ plasma. Controlling for the covariates age, gender, education, and diagnosis did not have a significant effect on any Pearson correlations. No correlations were improved by more than 0.10. The full table of correlations, including Pearson Rs and p-values, is found in **Supplementary Table 1**. The complete data set for the venous and Tasso+ plasma biomarker NPQ values is available in **Supplementary Table 2**.

**Figure 2.**
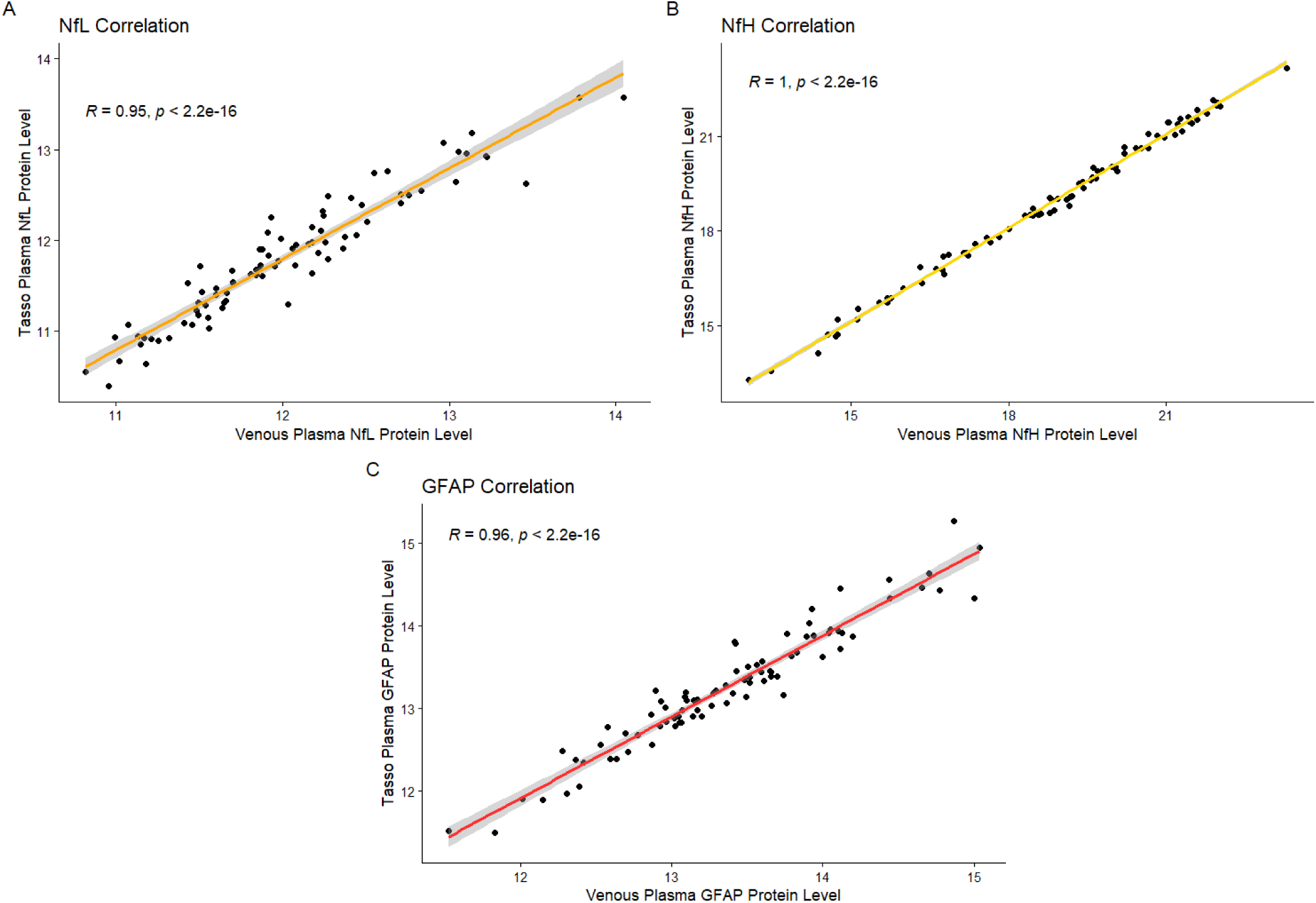
Biomarker ratio correlations between venous and Tasso+ plasma for other prominent neurodegenerative disease biomarkers: NfL (A), NfH (B) and GFAP (C). Lines represent linear regression results for Pearson’s R with 95% confidence intervals.

**Figure 3.**
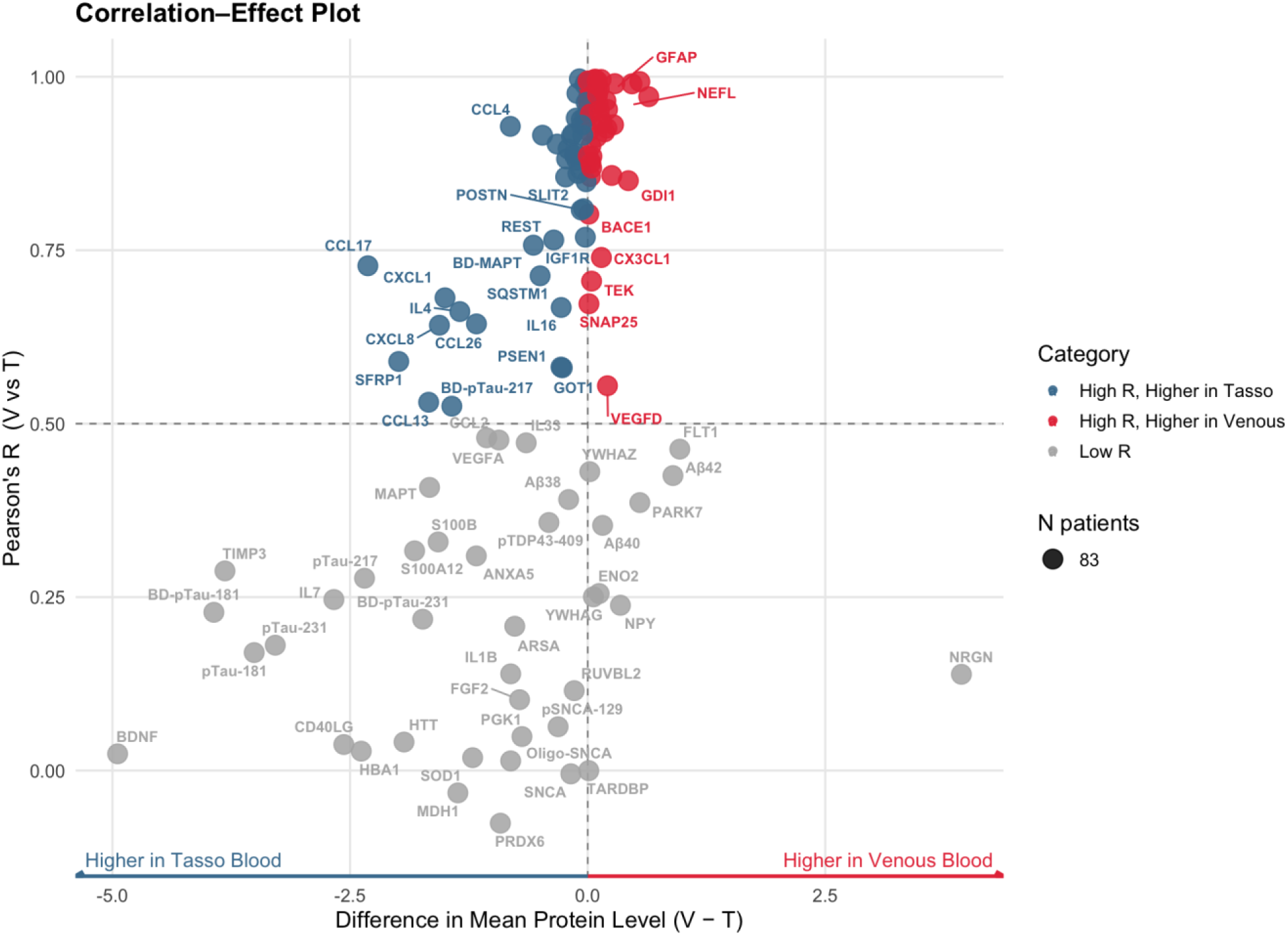
Correlation effect plot for all 131 biomarkers measured on the NULISA CNS120 panel. Red biomarkers have higher protein levels in venous blood, and blue biomarkers have higher protein levels in Tasso+ blood. Grey biomarkers have a low correlation coefficient.

### 3.5 APOE Correlation and Concordance with Genotyping

Of the 84 participants, 39 had apolipoprotein e4 genotype (*APOE e4*) status. ApoE4 was measured in both venous and Tasso+ plasma. ApoE4 showed an extremely strong correlation between the two plasma types (*R=*0.99, p<2.2×10^-16^). To determine concordance with genotyping, Tasso+ ApoE4 levels were compared to *APOE4* genotype status. If a participant had one or more copies of the *APOE4* allele, they were deemed positive. A Mann-Whitney U test between the two groups, *APOE4* positive and *APOE4* negative, yielded a p-value of 1.1×10^-6^, demonstrating a significant difference between the Tasso+ NPQ values for *APOE4* positive and negative individuals (**Supplementary Figure 1**).

## 4. Discussion

In this study, we evaluated the suitability of upper-arm capillary blood collected using the Tasso+ device for measuring well-established and explorative blood biomarkers for ADRD. We found that upper-arm capillary collection provides excellent agreement for many neurodegeneration-associated proteins in more than one-third of all proteins evaluated, with particularly strong concordance for NfL, NfH, GFAP, and *APOE* genotyping assessed on the protein level. In contrast, core AD biomarkers, including p-tau variants and amyloid-β peptides, showed substantially weaker agreement, although the performance of p-tau217 improved considerably when expressed as a ratio with p-tau181, particularly for BD-p-tau217.

The most important finding was that BD-p-tau217 improved the agreement between Tasso+ and venous plasma compared with conventional p-tau217, despite the systematic increase in p-tau concentrations observed in Tasso-derived samples, a finding that has been reported previously [22]. Importantly, we demonstrate for the first time that this upward shift in concentrations is also present for BD-p-tau217 and other BD-p-tau isoforms. More broadly, elevated NPQ values were observed for a substantial proportion of proteins measured in Tasso+ plasma relative to matched venous plasma, suggesting that this systematic increase extends beyond p-tau and may represent a general feature of upper-arm capillary blood collection. However, this was not universal, as biomarkers such as NfL, NfH, and GFAP showed excellent agreement between Tasso+ and venous plasma with little or no systematic difference in concentration.

Several mechanisms may be hypothesized as contributing to the systematically higher protein concentrations observed in Tasso-derived plasma. First, capillary blood differs physiologically from venous blood, comprising a mixture of arterial and venous blood together with small amounts of interstitial fluid. This unique composition may alter the concentration of circulating proteins. Second, the prolonged collection process, during which blood accumulates in the collection tube over several minutes, may permit localized tissue responses or subtle cellular activation that increases the release of intracellular or membrane-associated proteins into plasma. Third, differences in anticoagulant mixing, plasma recovery, or residual cellular material may result in slightly greater carryover of platelets, leukocytes, or extracellular vesicles, all of which are sources of CNS-associated proteins.

Notably, expressing BD-p-tau217 as a ratio with BD-p-tau181 largely compensated for these systematic differences, improving the correlation with venous plasma from *R=*0.53 for BD-p-tau217 alone to *R=*0.78 for BD-p-tau217/BD-p-tau181. This substantially outperformed conventional p-tau217 measured as a single biomarker (*R=*0.28).

By enabling minimally invasive, self-administered blood collection, Tasso+ carries great promise to lower the threshold for participation in research by improving access to biomarker testing in community and remote settings, among others. However, despite its ease of use, Tasso+ still requires timely sample processing and is likely to require cold-chain shipment to preserve biomarker integrity. As such, DBS sampling may retain important practical advantages if comparable analytical performance can be achieved. Rather than competing technologies, venipuncture, upper-arm capillary collection, and DBS are likely to have complementary roles, with the optimal sampling method depending on the intended application and setting.

Despite these promising results, several limitations should be acknowledged. First, despite highly statistically significant results, the sample size was relatively modest, and the study population was relatively homogenous and composed of mostly white older persons in the Phoenix metropolitan area receiving clinical care or participating in research studies at a specialized AD/ADRD institute. While supporting high internal validity, this may potentially limit the external validity of the results to a more ethno-racially diverse population of younger, older or middle-aged adults. Larger validation studies that are more representative of other populations of interest will therefore be required. Second, all Tasso+ samples in this study were collected and processed immediately in a clinical research setting with high proficiency for sample collection and processing. It will also be important to determine whether alternative processing workflows, including delayed processing, influence biomarker performance. Lastly, in Tasso+ collection, we only evaluated EDTA plasma and a single processing protocol, and it is possible that alternative collection tubes, preservative systems and centrifugation strategies may improve the stability and measurement of certain biomarkers in Tasso+ collection.

In conclusion, upper-arm capillary blood collected using the Tasso+ device shows considerable promise for measuring blood biomarkers of neurodegenerative disease. While several established AD biomarkers, particularly p-tau isoforms, exhibited systematic differences compared to venous plasma, BD-p-tau217 and ratio-based approaches substantially improved concordance to venous plasma measures. In contrast, biomarkers such as NfL and GFAP demonstrated excellent agreement between Tasso+ and venous plasma, supporting the utility of this collection method for a broad range of neurodegeneration-associated proteins. These findings provide an important foundation for the continued development of minimally invasive blood collection approaches and highlight both the opportunities and remaining challenges for translating blood-based biomarkers beyond specialist clinics and research settings.

## Data Availability

All data produced in the present work are contained in the supplementary tables within the manuscript.

## Acknowledgements

We are extremely grateful and indebted to the dedicated participants and their partners who graciously contributed to this study, without their altruistic support, engagement and participation this research would not be possible.

## Declaration of financial/other relationships

Alireza Atri: Researcher (paid to institution for contracted clinical trials): Alzheon, Athira, Biogen, Cognition Therapeutics, Eisai, Lilly, Vivoryon; Consultant/advisor (paid or unpaid): AriBio, Axsome, BMS, CoreEvitas, Eisai, Siemens Healthineers, Johnson & Johnson, Lantheus/Life Molecular Imaging, Lundbeck, Merck, Novo Nordisk, ONO, Prothena, Vaxxinity; and Book royalty: Oxford University Press. Jessica Langbaum reported receiving grants from the National Institute on Aging (NIA) and the Arizona Department of Health Services via the Arizona Alzheimer’s Consortium during the conduct of this study, Banner Health received institutional grants or contracts from Eli Lilly; she received personal fees from Premiere Inc. outside the submitted work. Nicholas J. Ashton reported receiving grants from the National Institute on Aging (NIA) and the Arizona Department of Health Services via the Arizona Alzheimer’s Consortium during the conduct of this study. He serves on scientific advisory boards for Alamar Biosciences (ADDF initiative), Biogen, New Amsterdam Pharma, Bristol Myers Squibb, and Abbott. He has received consultancy/speaker fees from Alamar Biosciences, Bioartic, Biogen, Eli-Lilly, Neurogen Biomarking, Roche, Spear Bio, Quanterix and Vigil Neurosciences. All other authors have nothing to disclose.

