## Supplementary Figure 1 for "Performance of upper-arm capillary blood collection for Alzheimer’s disease and central nervous system biomarkers: comparison of Tasso+ and venous plasma"

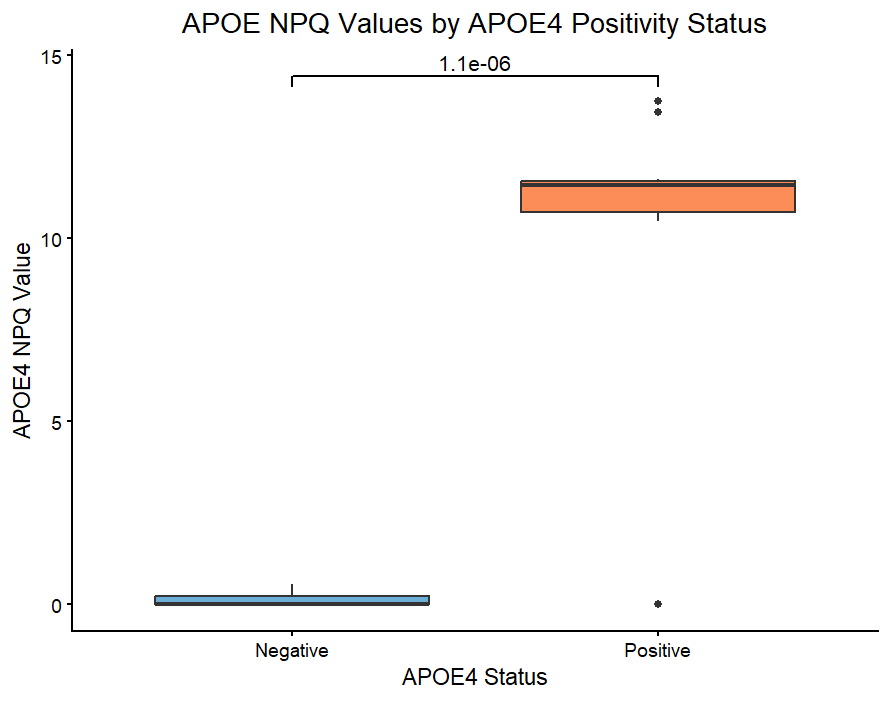

**Supplementary Figure 1.** ApoE4 status as predicted by Tasso+ plasma NPQ values. ApoE4 status was available for 39 out of 87 participants through past genotyping.
